# DNA methylation of Nuclear Factor of Activated T Cells 1 mediates the prospective relation between exposure to different traumatic event types and post-traumatic stress disorder

**DOI:** 10.1101/2021.11.17.21262624

**Authors:** James R. Occean, Agaz H. Wani, Janelle Donglasan, Allison E. Aiello, Sandro Galea, Karestan C. Koenen, Annie Qu, Derek E. Wildman, Monica Uddin

## Abstract

**Background:** The mechanisms through which exposure to differing trauma types become biologically embedded to shape the risk for subsequent post-traumatic stress disorder (PTSD) is unclear. DNA methylation (5-mC), particularly in stress-relevant genes, may play a role in this relationship.

**Methods:** We conducted path analysis using generalized structural equation modeling to investigate whether blood-derived 5-mC in Nuclear Factor of Activated T Cells 1 (*NFATC1*) mediated the prospective association between each of five different trauma types (“assaultive violence”, “other injury or shocking experience”, “learning of trauma to loved one”, “sudden, unexpected death of a close friend or relative”, and “other”) and lifetime PTSD assessed prospectively in the Detroit Neighborhood Health Study (n=183).

**Results:** All five trauma types were significantly associated with reduced methylation at *NFATC1* CpG site, cg17057218. Three of the five trauma types were significantly associated with increased methylation at *NFATC1* CpG site, cg22324981. Moreover, methylation at cg17057218 significantly mediated 23-34% of the total effect for three of the five trauma types (assaultive violence, other injury or shocking experience, and learning of trauma to a loved one), while methylation at cg22324981 mediated 36-53% of the total effect for two of the five trauma types (other injury or shocking experience and other). These CpG sites were differentially associated with transcription factor binding sites and chromatin state signatures.

**Limitations:** Prospective assessment of lifetime PTSD, rather than PTSD onset.

**Conclusions:** *NFATC1* 5-mC may be a potential mechanism in the relationship between some trauma types and prospective risk for PTSD. This finding may help inform treatment for trauma-specific PTSD.

## 1. Introduction

Post-traumatic stress disorder (PTSD) is a disabling and heterogeneous psychiatric disorder with an estimated lifetime prevalence of 6.8% among U.S. adults[1] and 3.9% globally[2]. PTSD is unique from other psychopathologies because its diagnostic criteria necessitate exposure to a traumatic event involving significant injury, actual or threatened death[3]. Exposure to trauma is thus an etiological component of PTSD. Yet, epidemiological reports indicate that the disorder will manifest in only a subset of trauma-exposed individuals[4]—suggesting the possibility of other factors that may shape the risk of PTSD development.

The psychological susceptibility or vulnerability to PTSD is related, at least in part, to the form or type of trauma exposure. Indeed, significant differences in conditional risk of PTSD associated with exposure to different trauma types are well documented. In a nationally representative study, Kessler *et al*. [5] reported an estimated conditional probability of PTSD ranging from 1.8% to 65.0% for men, and 5.4% to 48.5% for women, associated with exposure to specific traumatic events.

Among the study participants with PTSD, the events nominated as “most upsetting” were combat for men and rape for women[5]. Breslau *et al*.[6] later classified a list of 19 traumatic events into four composite classes and identified exposure to assaultive violence as conferring the highest conditional risk for PTSD. Similar findings were obtained from a predominantly African American sample by Goldmann *et al*.[7]; these authors reported experiences of assaultive violence conferred the highest risk for lifetime probable PTSD, followed by other injury or shock, sudden, unexpected death of a close relative or friend, and learning of traumas to a close relative or friend. Recent work from the WHO World Mental Health surveys has also shown a differential risk of PTSD across trauma types, specifically, an increased risk of PTSD and longer duration following exposure to violence-related traumas[4]. Collectively, these findings provide evidence of a relationship between exposure to different types of traumas and PTSD development, however, the mechanisms by which these exposures confer differing conditional risk for PTSD is unclear.

Biological insights into PTSD pathophysiology point to alterations of the hypothalamic-pituitary-adrenal axis, the body’s central stress response system that orchestrates the release of glucocorticoids (e.g., cortisol) following exposure to stress [8, 9]. Consequently, multiple studies have investigated epigenetic variations associated with PTSD development in stress-relevant pathways, such as the glucocorticoid receptor regulatory network (GRRN)[10–13]. The GRRN is active in peripheral tissues, including blood—one of the primary tissues that transduce the HPA axis response to stress throughout the body (reviewed in[14]). Previous work has shown that genes belonging to the GRRN pathway undergo epigenetic modifications following exposure to glucocorticoids such as cortisol[15]. Epigenetic mechanisms influence chromatin architecture and govern DNA accessibility to transcriptional machinery, thereby playing an important role in gene regulation (reviewed in[16]). As such, epigenetic modifications, particularly in GRRN genes, represent a potential mechanism wherein environmental exposures such as trauma may become biologically embedded to confer risk for PTSD. The most well-characterized epigenetic modification thus far in relation to PTSD, is DNA methylation (or 5-methylcytosine [5-mC]), which refers to the biochemical addition of a methyl (-CH_3_) group to the 5’ position of a cytosine base. Prior research has shown that 5-mC of GRRN genes, measured in blood, is associated with PTSD[13, 17], depression[18, 19], and is predictive of prospective risk for PTSD[20].

Concerning genes involved in the GRRN pathway, recent studies have implicated 5-mC in the gene encoding Nuclear Factor of Activated T Cells 1 (*NFATC1*) in PTSD[20, 21]. *NFATC1* (located on chromosome 18: 77,160,326-77,289,323) encodes a Ca^2+^/calmodulin-dependent isoform of the NFAT transcription factor family and is involved in several physiological processes (reviewed in[22]), including immune function regulation[23]. Differential methylation of immune-related genes is extensively documented in PTSD[24–27]. Moreover, recent work showed an altered transcriptional response of *NFATC1* following *in vitro* glucocorticoid stimulation[28], suggesting the gene plays a role in modulating stress response and may thus plausibly contribute to the development of stress-related psychopathology, including PTSD.

To date, however, only a few molecular studies have investigated trauma-related differences in PTSD. One study by Breen *et al*.[29] employed a combined transcriptome-wide peripheral blood study that included seven trauma types; they reported differences in gene expression signatures between interpersonal- and combat-related traumas that ultimately converged on shared biological processes. Another transcriptomic study by Huckins *et al*.[30] identified a gene associated with military, but not civilian, PTSD cohorts—indicating possible unique genetic etiologies in trauma-specific PTSD. An epigenome-wide association study (EWAS) of PTSD in civilians identified two genome-wide significant CpG sites in *NRG1* and *HGS*, respectively[31], which were not observed in subsequent EWAS analyses of military-derived cohorts[32], suggesting potential mechanistic differences, possibly in addition to other factors (e.g., sex), in the loci that mediate civilian- and military-related PTSD. Nevertheless, the molecular mechanisms underlying the relationship between exposure to different trauma types and risk for subsequent PTSD remains largely unclear, and an investigation of 5-mC in genes previously identified in stress-related psychopathology, of particular interest, *NFATC1*, may help elucidate such mechanisms.

To address this gap in knowledge, the present study sought to investigate the association between trauma types, *NFATC1* 5-mC, and lifetime PTSD. To this end, we focused our initial efforts on identifying PTSD-relevant CpG sites in *NFATC1.* We then interrogated the relation between different trauma types, methylation at the candidate CpG sites, and PTSD risk using a statistical mediation model. The different trauma types were hypothesized to confer differing conditional risk for prospective PTSD, mediated in part by potentially distinct methylation patterns of *NFATC1*.

## 2. Methods

### 2.1. Study participants

Samples included in this study were drawn from the Detroit Neighborhood Health Study (DNHS), a longitudinal population-based study composed of predominantly African American adult residents in Detroit, Michigan. The study was approved by the institutional review boards of the University of Michigan and the University of North Carolina at Chapel Hill. Details regarding the DNHS methodology have been described elsewhere[7, 27]. Briefly, after written informed consent was obtained, the DNHS collected self-reported data on participant demographics (i.e., age, gender, and race), trauma exposures, and PTSD symptoms. Informed consent was again obtained for participants who elected to provide biospecimen.

The present investigation was limited to trauma-exposed individuals with available peripheral blood-derived genomic DNA (gDNA) samples from wave 1 (W1) or wave 2 (W2) of the DNHS. Participants with gDNA samples from W1 were included only if they reported no trauma exposure in W2 or if the W2 trauma exposure fell within the same category as the W1 trauma (trauma types are described below).

### 2.2. Measures

#### 2.2.1. W3 lifetime PTSD

Participants were provided with a list of 19 traumatic events and asked to select the event(s) that they had experienced in their lifetime or since the last interview for post-baseline waves. From the experienced events, the respondent then nominated the “worst” event. In baseline waves, a random event was also selected for respondents who experienced multiple traumatic events. PTSD symptoms were assessed in reference to both the nominated worst and the randomly selected event using the civilian version of the PTSD checklist (PCL-C)[33] and supplemented additional questions regarding duration, timing, and impairment due to symptoms. The respondents were considered affected with lifetime PTSD in wave 3 (W3) if they met all *Diagnostic and Statistical Manual of Mental Disorders* (4th ed.; *DSM-IV*)[34] criteria in reference to either the worst or randomly chosen traumatic event in W1, W2, or in relation to a new worst trauma that may have occurred between W2 and W3. Missing data for W3 lifetime PTSD (n=39) were imputed using the *mice* package[35] in R. Data inspection with density plots showed similar distribution between imputed and observed data, which indicates imputed data are indeed plausible values. The PTSD instrument described in this study was validated in the DNHS by blinded clinical interviews of a random subsample and yielded excellent internal consistency and high concordance[36].

#### 2.2.2. W1/W2 5-mC

As previously mentioned, this study included participants who consented to provide a biospecimen, specifically, peripheral blood drawn via venipuncture by a phlebotomist. Detailed methods regarding sample processing in the DNHS have been described previously[27, 37]. Briefly, gDNA was isolated from peripheral blood using the Qiagen DNA Mini Kit (Qiagen, Hilden, Germany), bisulfite-converted using the Zymo EZ-96 DNA methylation kit (Zymo Research, Irvine, CA), then methylation tested using the Illumina Infinium Human Methylation 450K BeadChip platform (Illumina, San Diego, CA), as previously reported[38, 39]. All steps were completed following the manufacturer’s recommended protocol.

A battery of quality control (QC) steps was performed on the resulting 5-mC data. First, to account for potential sex bias associated with methylation data, samples were assessed for sex discordance using the *minfi*[40] R package and removed if the participant’s self-reported gender differed from the predicted sex. QC was next performed on the raw 5-mC beta values obtained via the Illumina GenomeStudio software and filtered for poorly performing samples, i.e., those with low mean signal intensity (<2000 arbitrary units or <50% of the overall median)[41] or >10% missing value. Probes with detection *p*-value >0.01 were set to missing and those with >10% missing values were removed along with cross-reactive and polymorphic probes[42]. To correct for the type II probe bias, 5-mC data was normalized using the beta-mixture quantile method[43–45]. Finally, *ComBat* adjustment was employed on logit-transformed beta-values (M-values), while controlling for trauma types, W3 lifetime PTSD, and sex, to account for potential batch effects via the empirical Bayes method[46] used in the *sva*[47] R package. *ComBat* adjustment requires no missing data, thus, probes with missing values were imputed using the K-nearest neighbor algorithm[48] then set to missing after *ComBat* adjustment. All participants included in this study had 5-mC measures at only one time-point (W1/W2).

#### 2.2.3. W1/W2 trauma types

The main exposure variable of interest was the W1/W2 trauma types. Study participants were asked about traumatic experiences in their lifetime (in baseline waves) and since their last interview (in post-baseline waves; W2) from a list of 19 traumatic events. These events were categorized into four traumatic event types: “assaultive violence”, “other injury or shocking experience”, “learning of trauma to a loved one”, and “sudden, unexpected death of a close friend or relative”, as previously described [6, 7]. An additional “other” category was included for respondents who reported experiencing “other extraordinarily stressful situation or event”. Since W3 lifetime PTSD was assessed in reference to both the worst event nominated by the respondent and a randomly selected event, each trauma type variable reflects either the worst event nominated by the participant in W1/W2 or the baseline randomly selected event. Our study did not include traumatic events from W3 as these experiences would have occurred after W1/W2 methylation assessments and conflicted with the *a priori* hypothesized prospective relationship between each trauma type, 5-mC, and subsequent PTSD.

#### 2.2.4. Covariates

The present study included sex, age, ancestry, and cell proportions as covariates. Genomic ancestry estimates were derived from a principle component-based approach involving methylation probes near single-nucleotide polymorphisms [49]. Principal components two to four were included in this study as ancestry components to account for population stratification, as previously performed[50]. Lastly, cell proportions were computed using the algorithm developed by Houseman [51] implemented in the *EpiDISH*[52, 53] R package. We included estimates of NK cells, monocytes, B cells, CD8+ T cells, and CD4+ T cells as covariates for cell proportions.

### 2.3. Statistical analyses

Linear regression models were implemented using the *CpGassoc*[41] R package to identify individual *NFATC1* CpG sites associated with W3 lifetime PTSD while controlling for other study covariates. Additionally, *post hoc* analysis was conducted using a Wilcoxon rank sum test to assess differences in methylation levels at candidate CpG sites between participants with and without W3 lifetime PTSD. Here, *p*-values were corrected for multiple hypothesis testing using the Benjamini-Hochberg[54] method (false discovery rate [FDR] < 0.05) and the CpG sites significantly associated with W3 lifetime PTSD were retained for downstream analyses. All analyses mentioned above were performed in R v3.6.0[55].

The association between trauma types, candidate *NFATC1* CpG sites, and W3 lifetime PTSD were assessed using path analysis implemented in a generalized structural equation model (GSEM). Because W3 lifetime PTSD is a binary outcome variable, the GSEM was carried out using the Bernoulli-logit link function in Stata v15.1[56]. GSEM was the method used to assess this relationship because it simultaneously tests the relationships among study variables, which reduces the chance of inflating the Type I error from multiple hypothesis testing[57]. Furthermore, to assess potential mediation effects, the average direct effect (DE), indirect effect (IE), and total effect (TE) along with mediated proportion and 95% confidence intervals (CI) were estimated with 1000 simulations for each trauma type and candidate *NFATC1* CpG sites, while adjusting for covariates, using the *medeff* command in the *mediation*[58] Stata package. The associations and effects between trauma types, candidate *NFATC1* CpG sites, and W3 lifetime PTSD were considered statistically significant at a nominal *p*-value of 0.05 and when lower and upper 95% CIs did not overlap the null (i.e., zero) value.

### 2.4. Potential functional relevance of NFATC1 5-mC

To assess the potential functional relevance of the CpG sites that were identified as significant mediators between a trauma type and prospective risk for PTSD, the CpG sites were annotated to transcription factor binding sites (TFBS) and chromatin state signatures using the publicly available ENCODE 3 TFBS track[59] and Broad ChromHMM track[60], respectively, from the hg19 UCSC Genome Browser[61]. The ENCODE 3 TFBS track was generated by the ENCODE Consortium and consists of an extensive set of human TFBS identified from ChIP-seq experiments. For the Broad Institute’s ChromHMM track, a multivariate Hidden Markov Model was used to segment the genome into 15 chromatin states representing the combinatorial pattern of nine chromatin marks from ChIP-seq experiments—distinctions in the 15 defined chromatin states were also functionally validated[60]. In line with other genomic studies of whole blood[62], we used the GM12878 B-lymphoblastoid cell line for all functional annotations as it is the most similar available cell type to whole blood. Lastly, to place our blood-based findings in the context of the brain, the tissue of interest for psychiatric disorders, we accessed publicly available data from the Human Protein Atlas (http://www.proteinatlas.org) and assessed the regional specificity and distribution of *NFATC1*’s expression levels in the brain[63]. This database contains gene consensus normalized expression levels, summarized from the GTEx (Genotype-Tissue Expression) project and the FANTOM5 dataset, for 10 brain regions (i.e., cerebral cortex, olfactory region, hippocampal formation, amygdala, basal ganglia, thalamus, hypothalamus, midbrain, pons and medulla, and cerebellum)[63].

## 3. Results

### 3.1. Sample characteristics

Our study included 183 trauma-exposed participants with available gDNA samples from W1/W2 of the DNHS. Among these participants, 106 (57.9%) met the criteria for lifetime PTSD in W3. The demographic characteristics of the participants and the type of trauma exposures are described in Table 1. This study consisted of mostly female (*n*=117; 63.9%) and Black (*n*=154; 84.2%) participants with a mean age of 53.8. The most and least frequently nominated “worst” or randomly selected trauma types were other injury or shocking experience (*n*=91; 49.7%) and other (*n*=40; 21.9%), respectively. Of the trauma types assessed, the participants exposed to the “other” trauma type (versus participants who did not) had the highest risk for W3 lifetime PTSD (*n*=28; 70.0%) followed by exposure to assaultive violence (*n*=50; 66.7%).

**Table 1.**
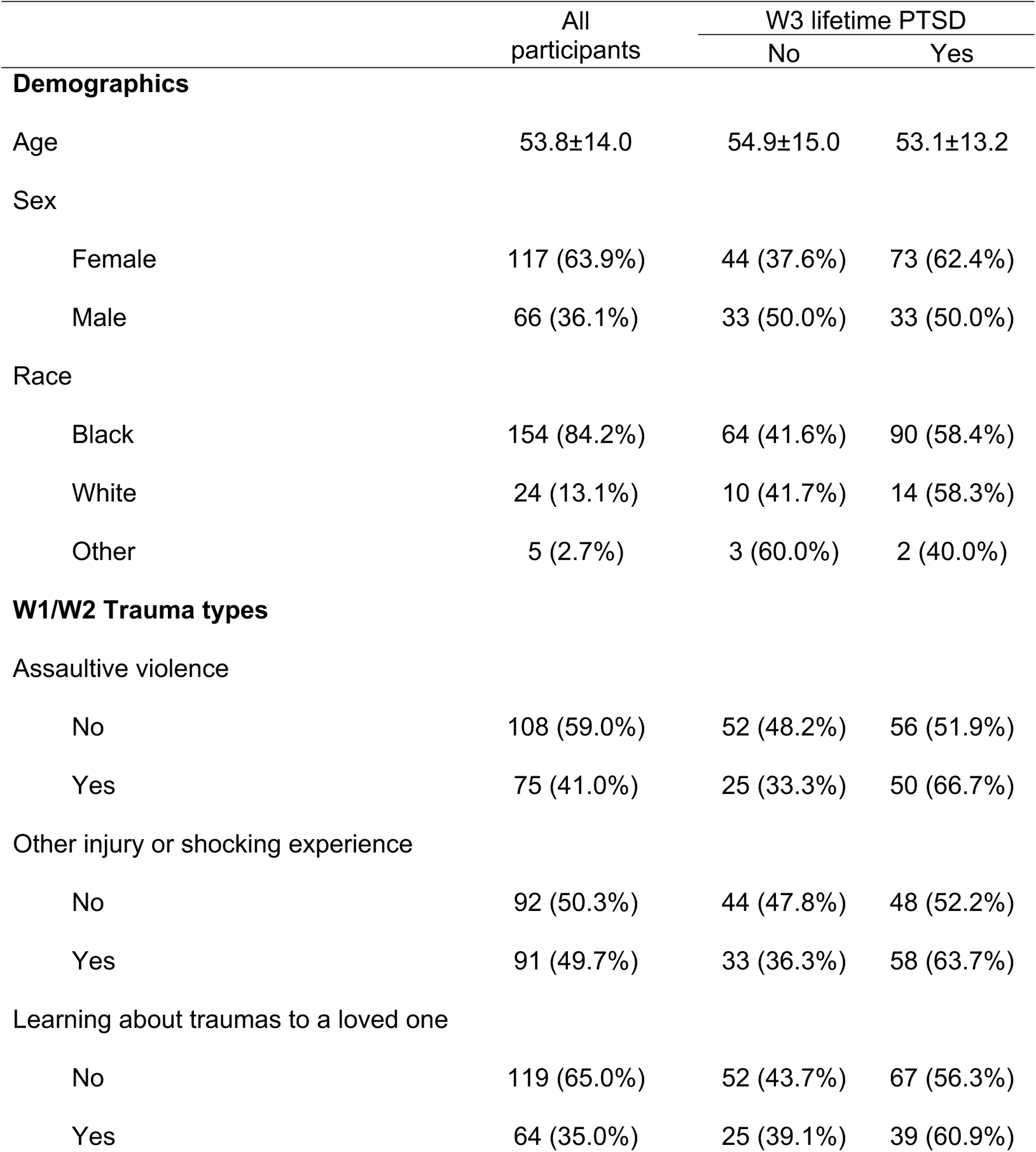
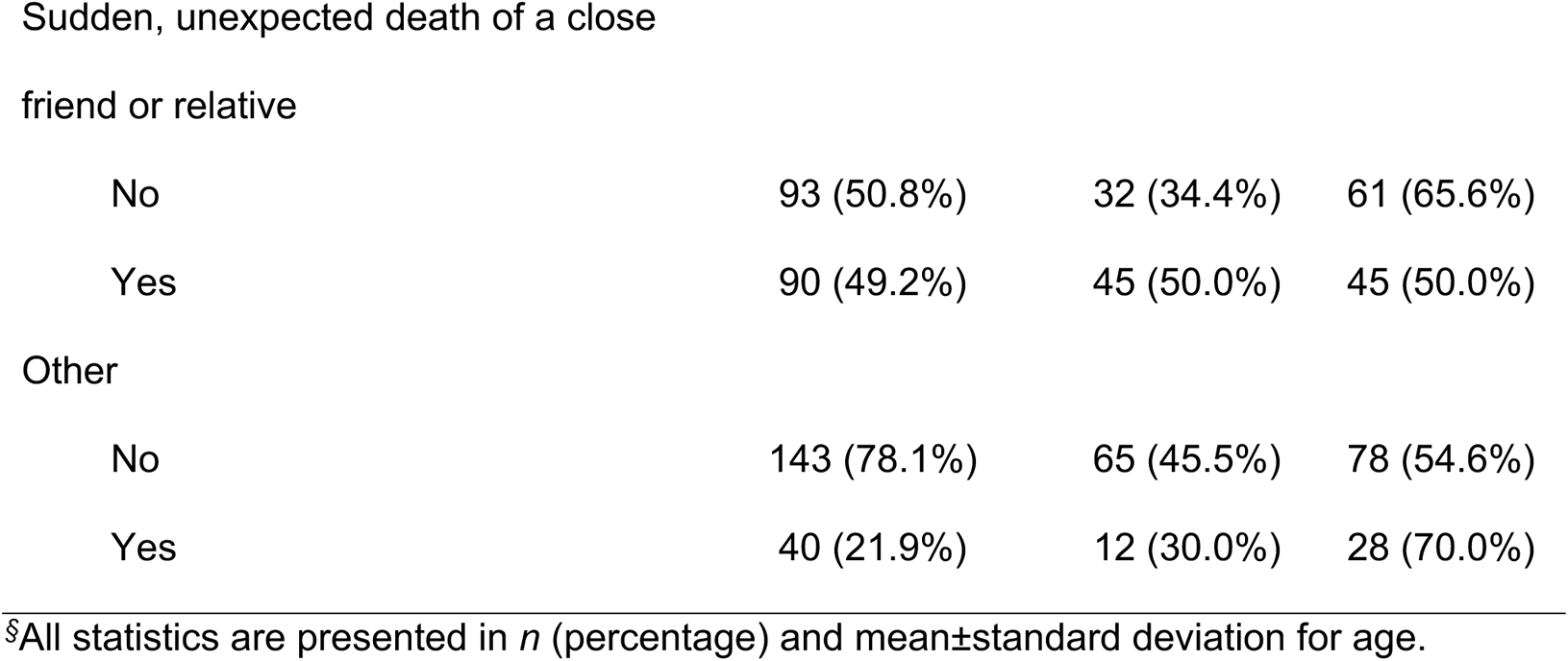
Sample characteristics, *n*=183^§^

### 3.2. NFATC1 CpG sites associated with W3 lifetime PTSD

A total of 160 out of 166 methylation microarray probes associated with *NFATC1* survived QC assessment. The relationship between *NFATC1* 5-mC and W3 lifetime PTSD was assessed at the 160 CpG sites while controlling for study covariates. Among the 160 CpG sites, 26 were nominally associated with W3 lifetime PTSD. The top ten CpG sites are reported in Table 2, along with the annotations provided by Illumina, F-statistic, and the corresponding nominal and FDR-corrected *p*-value. After correction for multiple hypothesis testing, three CpG sites remained significantly associated with W3 lifetime PTSD: cg17057218 (F=15.31, *p* <0.001, FDR=0.021), cg04755409 (F=13.78, *p* <0.001, FDR=0.022), and cg22324981 (F=12.99, *p* <0.001, FDR=0.022). Genomic annotations showed cg17057218 was positioned within the gene body, 5’ UTR, and CpG shelf, cg04755409 was positioned within the gene body and CpG island, and cg22324981 was positioned within the gene body and CpG shore. Further interrogations of cg17057218, cg04755409, and cg22324981 in *post hoc* analysis supported the finding of significant differences in 5-mC levels between participants with and without W3 lifetime PTSD (Figure 1). Specifically, those that met the criteria for lifetime PTSD in W3 had significantly lower methylation levels in W1/W2 at cg17057218 (W=5228, *p*=0.001, FDR=0.002) and cg04755409 (W=5345, *p* <0.001, FDR=0.001), but significantly higher methylation levels at cg22324981 (W=3240, *p*=0.018, FDR=0.018).

**Figure 1.**
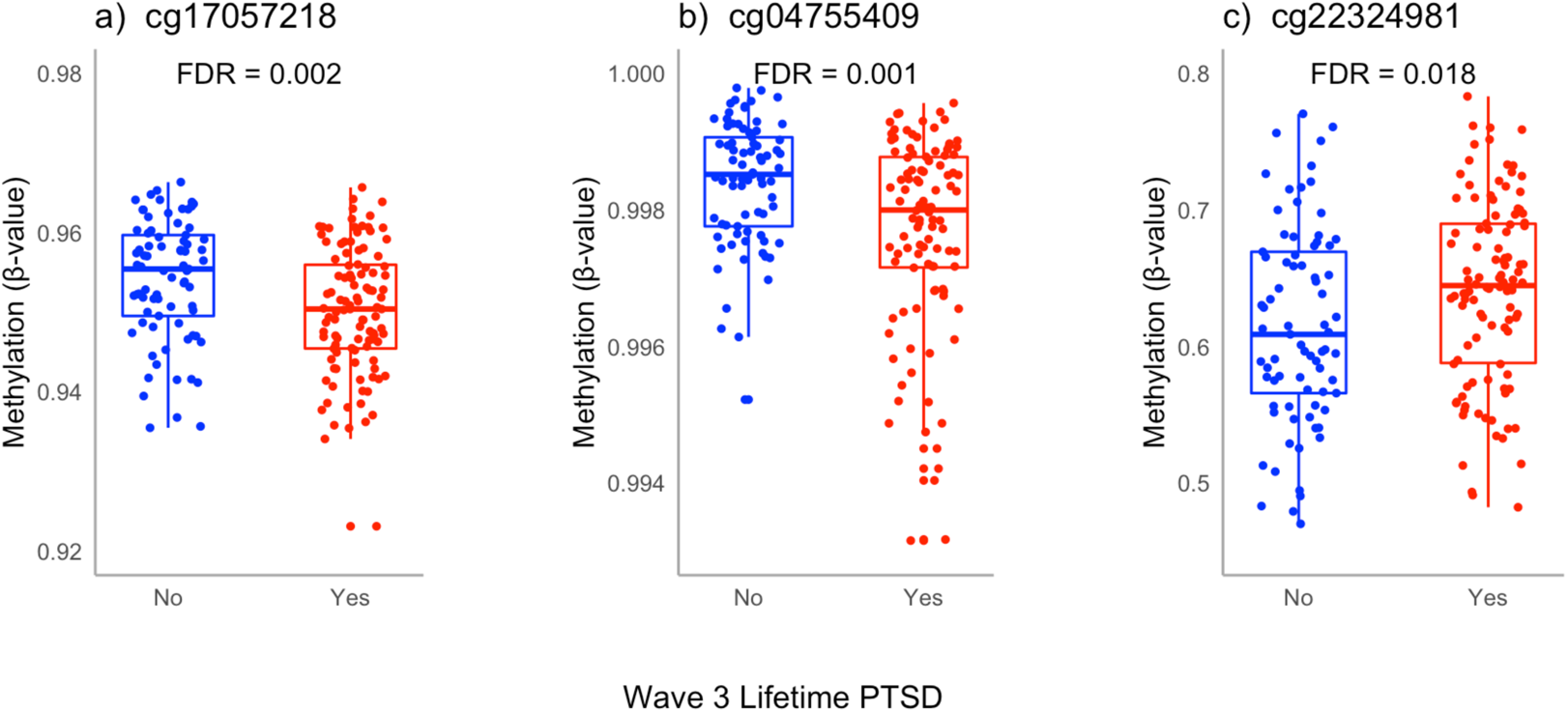
Methylation at cg17057218, cg04755409, and cg22324981 between participants with and without W3 lifetime PTSD. Participants with W3 lifetime PTSD had lower W1/W2 methylation at cg17057218 (W=5228, *p*=0.001, FDR=0.002) and cg04755409 (W=5345, *p* <0.001, FDR=0.001), but higher methylation at cg22324981 (W=3240, *p*=0.018, FDR=0.018) based on Wilcoxon rank sum test.

**Table 2.** Characteristics of the top 10 *NFATC1* CpG sites associated with W3 lifetime PTSD^§^

| Illumina ID (genomic coordinates) | Genomic annotation | CpG annotation | F-statistic | p-value | FDR |
| --- | --- | --- | --- | --- | --- |
| cg17057218 (Chr18:77,183,423) | Body;5'UTR | Shelf | 15.31 | 1.32E<sup>-04</sup> | **0.021** |
| cg04755409 (Chr18:77,233,327) | Body | Island | 13.78 | 2.77E<sup>-04</sup> | **0.022** |
| cg22324981 (Chr18:77,283,493) | Body | Shore | 12.99 | 4.09E<sup>-04</sup> | **0.022** |
| cg05718035 (Chr18:77,240,530) | Body | Island | 9.80 | 2.05E<sup>-03</sup> | 0.082 |
| cg00431336 (Chr18:77,223,422) | Body | Shore | 9.17 | 2.84E<sup>-03</sup> | 0.091 |
| cg07952877 (Chr18:77,171,705) | Body;5'UTR | Island | 8.34 | 4.38E<sup>-03</sup> | 0.115 |
| cg15784814 (Chr18:77,171,832) | Body;5'UTR | Island | 8.07 | 5.05E<sup>-03</sup> | 0.115 |
| cg26550337 (Chr18:77,203,542) | Body;5'UTR | Island | 6.60 | 1.10E<sup>-02</sup> | 0.221 |
| cg02743589 (Chr18:77,217,348) | Body | Shore | 6.13 | 1.43E<sup>-02</sup> | 0.227 |
| cg26100137 (Chr18:77,203,667) | Body;5'UTR | Island | 6.13 | 1.43E<sup>-02</sup> | 0.227 |

<sup>§</sup>Genomic annotation and CpG annotation (relation to UCSC CpG island) were determined from the annotation provided for each probe by Illumina. Coordinates and annotations are provided based on GRCh37/hg19 genome assembly.

### 3.3. Association between trauma types, methylation at cg17057218, cg04755409, and cg22324981, and W3 lifetime PTSD

In path analysis using GSEM, we investigated the association between each of five trauma types, methylation at cg17057218, cg04755409, and cg22324981, and W3 lifetime PTSD. The GSEM along with unstandardized coefficients is shown in Figure 2. All five trauma types were significantly associated with cg17057218 methylation and were consistent in showing a negative direction of effect: assaultive violence (B=-0.0048, *p*=0.002, 95% CI=[−0.0079, −0.0017]), other injury or shocking experience (B=-0.0050, *p*=0.001, 95% CI=[−0.0080, −0.0020]), learning of trauma to a loved one (B=-0.0048, *p*=0.002, 95% CI=[−0.0078, −0.0018]), sudden, unexpected death of a close friend or relative (B=-0.0044, *p*=0.004, 95% CI=[−0.0074, −0.0014]), and other (B=-0.0045, *p*=0.007, 95% CI=[−0.0077, −0.0012]) were all associated with reduced methylation at cg17057218. In contrast, three trauma types were significantly associated with cg22324981 methylation and were consistent in showing a positive direction of effect: other injury or shocking experience (B=0.0219, *p* <0.001, 95% CI=[0.0089, 0.0349]), sudden, unexpected death of a close friend or relative (B=0.0149, *p*=0.024, 95% CI=[0.0019, 0.0278]), and other (B=0.0262 *p* <0.001, 95% CI=[0.0122, 0.0403]) were all associated with increased methylation at cg22324981. No trauma types were significantly associated with methylation at cg04755409; thus, DE, IDE, and TE were not estimated for this CpG site.

**Figure 2.**
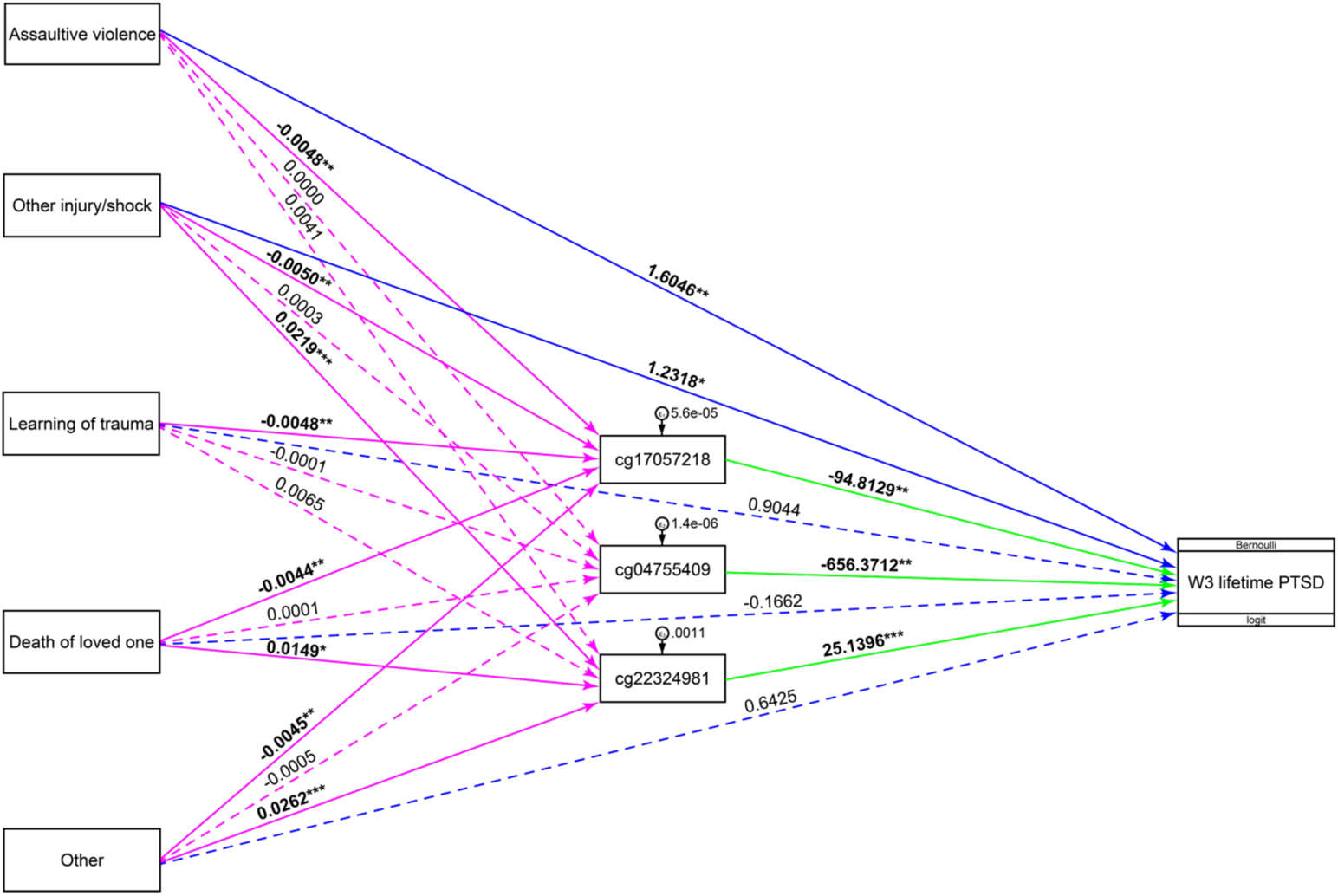
Path analysis using generalized structural equation modeling. Shown are unstandardized coefficients of exposures to mediators (magenta), mediators to outcome (green), and exposures to outcome (blue). Trauma types are abbreviated, and model is adjusted for sex, age, ancestry, and cell proportions (paths are not shown). Non-significant paths are dashed; \**p* <0.05; \*\**p* <0.01; \*\*\**p* <0.001.

In terms of prospective risk for PTSD, we observed significant negative associations between cg17057218 methylation and W3 lifetime PTSD (B=-94.8129, *p*=0.001, 95% CI=[−151.8257, −37.8000]), and cg04755409 and W3 lifetime PTSD (B=-656.3712, *p*=0.002, 95% CI=[−1061.9910, −250.7508]), but significant positive associations between cg22324981 and W3 lifetime PTSD (B=25.1396, *p* <0.001, 95% CI=[11.5744, 38.7048]). We also observed significant positive associations between assaultive violence and W3 lifetime PTSD (B=1.6046, *p*=0.009, 95% CI=[0.4080, 2.8011]); and other injury or shocking experience and W3 lifetime PTSD (B=1.2318, *p*=0.038, 95% CI=[0.0686, 2.3949]). No significant associations were observed between learning of trauma to a loved one and W3 lifetime PTSD, sudden, unexpected death of a close friend or relative and W3 lifetime PTSD, or other and W3 lifetime PTSD in the described GSEM.

### 3.4. Mediational role of methylation at cg17057218 and cg22324981 in the relationship between trauma types and W3 lifetime PTSD

To evaluate the mediational role of cg17057218 and cg22324981 methylation as a potential link between different trauma types and differential risk for PTSD, we estimated the DE, IDE, and TE. Table 3 reports the effects for the trauma types that were significantly associated with methylation at cg22324981 and/or cg17057218 from the path analysis GSEM.

**Table 3.** Effects of W1/W2 trauma types on W3 lifetime PTSD, as mediated by methylation at cg17057218 and cg22324981

|  | Direct effect<br>Mean [95% CI] | Indirect effect<br>Mean [95% CI] | Total effect<br>Mean [95% CI] | Proportion Mediated<br>[95% CI] |
| --- | --- | --- | --- | --- |
| cg17057218 |  |  |  |  |
| Assaultive violence | **0.2122**<br>[0.0567,0.3460] | **0.0642**<br>[0.0158,0.1302] | **0.2764**<br>[0.1290,0.4017] | **0.2292**<br>[0.1597,0.4975] |
| Other injury/shock | **0.1669**<br>[0.0064,0.3082] | **0.0841**<br>[0.0293,0.1560] | **0.2509**<br>[0.0998,0.3844] | **0.3311**<br>[0.2181,0.8352] |
| Learning of trauma | 0.1173<br>[-0.0416,0.2537] | **0.0643**<br>[0.0168,0.1267] | **0.1816**<br>[0.0303,0.3117] | **0.3437**<br>[0.1966, 1.5290] |
| Death of loved one | -0.0294<br>[-0.1848,0.1227] | **0.0691**<br>[0.0193,0.1332] | 0.0397<br>[0.1106,0.1790] | 0.6752<br>[-12.0433,20.5141] |
| Other | 0.0819<br>[-0.0928,0.2414] | **0.0682**<br>[0.0171,0.1380] | 0.1500<br>[0.0226,0.3018] | 0.4130<br>[-2.8303, 3.5310] |
| cg22324981 |  |  |  |  |
| Other injury/shock | **0.1680**<br>[0.0067,0.3125] | **0.0943**<br>[0.0336,0.1651] | **0.2623**<br>[0.0998,0.4060] | **0.3565**<br>[0.2317,0.9098] |
| Death of loved one | -0.0294<br>[-0.1852,0.1183] | **0.0661**<br>[0.0164,0.1273] | 0.0368<br>[0.1271,0.1851] | 0.5921<br>[-12.8998,13.1124] |
| Other | 0.0803<br>[-0.0937,0.2350] | **0.0991**<br>[0.0352,0.1719] | **0.1794**<br>[0.0016,0.3297] | **0.5304**<br>[0.2654,2.8677] |

Note: Trauma types are abbreviated. Significant effects (when lower and upper CIs do not overlap zero) are bolded.

#### 3.4.1. Mediation by cg17057218 methylation

A proportion of the TE for three of the five trauma types on W3 lifetime PTSD was significantly mediated by methylation at cg17057218. Specifically, methylation at cg17057218 significantly mediated 34% of the TE of learning of trauma to a loved one on W3 lifetime PTSD (mean_DE_=0.1173, 95% CI=[−0.0416, 0.2537]; mean_IDE_=0.0643, 95% CI=[0.0168, 0.1267]; mean_TE_=0.1816, 95% CI=[0.0303, 0.3117]), 33% of the TE of other injury or shocking experience on W3 lifetime PTSD (mean_DE_=0.1669, 95% CI=[0.0064, 0.3082]; mean_IDE_=0.0841, 95% CI=[0.0293, 0.1560]; mean_TE_=0.2509, 95% CI=[0.0998, 0.3844]), and 23% of the TE of assaultive violence on W3 lifetime PTSD (mean_DE_=0.2122, 95% CI=[0.0567, 0.3460]; mean_IDE_=0.0642, 95% CI=[0.0158, 0.1302]; mean_TE_=0.2764, 95% CI=[0.1290, 0.4017]). Methylation at cg17057218 did not significantly mediate the TE of sudden, unexpected death of a close friend or relative or other on W3 lifetime PTSD.

#### 3.4.2. Mediation by cg22324981 methylation

A proportion of the TE of two of the five trauma types on W3 lifetime PTSD was significantly mediated by methylation at cg22324981. Specifically, methylation at cg22324981 mediated 53% of the TE of other on W3 lifetime PTSD (mean_DE_=0.0803, 95% CI= [−0.0937, 0.2350]; mean_IDE_=0.0991, 95% CI=[0.0352, 0.1719]; mean_TE_=0.1794, 95% CI=[0.0016,0.3297]) and 36% of the TE of other injury or shocking experience on W3 lifetime PTSD (mean_DE_=0.1680, 95% CI=[0.0067, 0.3125]; mean_IDE_=0.0943, 95% CI=[0.0336, 0.1651]; mean_TE_=0.2623, 95% CI=[0.0998, 0.4060]). Methylation at cg22324981 did not significantly mediate the TE of assaultive violence, learning of trauma to a loved one, or sudden, unexpected death of a close friend or relative on W3 lifetime PTSD.

### 3.5. Potential functional and cross-tissue relevance of mediating CpG sites

Using publicly available data, we attempted to assess the functional relevance of cg17057218 and cg22324981. The ENCODE 3 TFBS track[59] showed no regional overlaps between cg17057218 and TFBS, while cg22324981 was shown to overlap several TFBS (i.e., ASH2L, BCL11A, EBF1, EP300, PAX5, and RBBP5). The Broad ChromHMM track[60] revealed that cg17057218 overlaps a weakly transcribed genomic region, whereas cg22324981 was shown to overlap a region denoting strong enhancer activity. Finally, we attempted to assess the cross-tissue relevance of *NFATC1* expression in the tissue of interest, the brain, using the Human Protein Atlas (http://www.proteinatlas.org)[63]. The data showed *NFATC1* was expressed in all 10 available brain anatomical regions (i.e., cerebral cortex, olfactory region, hippocampal formation, amygdala, basal ganglia, thalamus, hypothalamus, midbrain, pons and medulla, and cerebellum) and showed no brain regional specificity.

## 4. Discussion

Existing evidence suggests differing conditional risk for PTSD associated with the type of trauma exposure. The mechanisms underlying this relationship remain elusive and epigenetic modifications, particularly in stress-relevant pathways, may play a role. In this study, we investigated whether blood-derived 5-mC of *NFATC1,* a gene involved in the GRRN pathway, mediated the association between five different trauma types and prospective risk for lifetime PTSD. We observed that methylation at cg17057218 mediated a significant proportion of the TE on W3 lifetime PTSD for three of the five trauma types, while methylation at cg22324981 mediated a significant proportion of the TE for two of the five trauma types. Collectively, these results highlight a complex relationship between trauma types and 5-mC and suggest *NFATC1* 5-mC as a potential mechanism that partially mediates the relationship, between some (assaultive violence, other injury or shocking experience, learning of trauma to a loved one, and other), but not all, trauma types and prospective risk for PTSD.

This study reports several notable findings. Importantly, methylation at cg17057218 partially mediated the relations between three of five trauma types (assaultive violence, other injury or shocking experience, and learning of trauma to a loved one) and W3 lifetime PTSD. This finding was intriguing considering that two of the three trauma types (assaultive violence and other injury or shocking experience) can be broadly classified as directly experienced traumas. Such directly experienced trauma types have been associated with a relatively higher risk of PTSD and symptom presentation in comparison to indirectly experienced traumas[64]. Nevertheless, learning of trauma to a loved one, which could be broadly classified as an indirectly experienced trauma, was also significantly mediated by methylation at cg17057218. This suggests that qualitatively different trauma types, possibly beyond the context of direct and indirect traumatic events, may impact methylation at this locus. Furthermore, a differing pattern was observed for methylation at cg22324981. This CpG site partially mediated the relations between two trauma types (other injury or shocking experience and other) and W3 lifetime PTSD. Because the “other” trauma type can encompass several different trauma types, it is not possible to classify these two trauma types into an informative group. However, it is noteworthy that assaultive violence was not significantly associated with or mediated by cg22324981 methylation. One possible explanation is that while both assaultive violence and other injury or shocking experience can be classified as directly experienced trauma types, these trauma types may also have important and possibly differing biological relevance. Indeed, assaultive violence includes military combat exposure, whereas other injury or shocking experience includes civilian-related trauma types (e.g., natural disasters), and both have been contrasted in other biological studies of PTSD[29, 30]. In sum, these findings underscore the complex nature of trauma types and their potential molecular underpinnings.

Secondly, we observed a differing, but consistent, relationship between some of the trauma types and methylation patterns at cg17057218 and cg22324981. We observed that exposure to each of the five trauma types was associated with reduced methylation at cg17057218, which concurrently, was significantly associated with lifetime PTSD in W3. In contrast, three of the five trauma types were associated with increased methylation at cg22324981, which concurrently, was also significantly associated with W3 lifetime PTSD. Although not specific to *NFATC1*, previous studies of PTSD have reported both lower[26, 31] and higher[65] DNA methylation among persons with the disorder. Interestingly, the differing methylation patterns observed here between cg17057218 and cg22324981 seem consistent with their potential biological relevance. For instance, 5-mC is known to alter the binding of transcription factors to DNA (reviewed in[66]), and using publicly available data of TFBS and chromatin states characterized from a B-lymphoblastoid cell line, we observed an overlap between several transcription factors and cg22324981, whereas no overlaps were observed for cg17057218. Similarly, cg17057218 was shown to be positioned in a weakly transcribed genomic region, whereas cg22324981 was shown to be positioned within a region denoting strong enhancer activity. Thus, it seems plausible that the PTSD- and trauma-relevant 5-mC patterns observed here may be genomic context-specific.

Nevertheless, methylation at both CpG sites showed a consistent direction of relationship (negative for cg17057218 and positive for cg22324981) with their associated trauma types, and lifetime PTSD in path analysis, suggesting they may play a role in modulating stress response and related psychopathology, including PTSD. Indeed, in a secondary analysis of DNHS gene expression data[67] from a subset of individuals analyzed in this work (n=81), we observed a positive correlation between 5-mC and gene expression among those with lifetime PTSD and a negative correlation among trauma-exposed controls. Although neither correlation achieved statistical significance, these opposite directions of effect suggest that *NFATC1* may be differentially regulated among those with vs. without lifetime PTSD, as has previously been observed for other loci[31].

Overall, we observed that 23%-53% of the TEs for four of the five trauma types (assaultive violence, other injury or shocking experience, learning of trauma to a loved one, and other) was significantly mediated by *NFATC1* methylation. This observation coupled with recent evidence of altered gene expression of *NFATC1* following *in vitro* stimulation by the synthetic glucocorticoid, dexamethasone[28], and publicly available transcriptomics data[63] showing expression of *NFATC1* in the brain, further implicates this locus in stress-related psychopathology. These findings could also suggest that the response to physiological triggers (e.g., synthetic glucocorticoid) may similarly reflect exposure to contextually different trauma types, at least for *NFATC1*.

Although we found significant differences in methylation levels at another *NFATC1* CpG site, cg04755409, between participants with and without W3 lifetime PTSD, cg04755409 was not significantly associated with any of the five trauma types. Notably, methylation at this CpG site had the strongest absolute effect on W3 lifetime PTSD in the path analysis GSEM. Using publicly available data from IMAGE-CpG[68], a web-based search tool with correlational data between blood and brain 5-mC, we also observed a significant correlation between blood and brain methylation at cg04755409 (rho=0.76, *p*=0.016). Thus, it remains plausible that methylation at this CpG site may play a role in PTSD-related psychopathology, despite not being significantly associated with the trauma types assessed in this study.

Lastly, we found that exposure to other extraordinarily stressful situation or event conferred the highest risk for lifetime PTSD, followed by assaultive violence. Although the nature of the “other” trauma type is unknown, this finding of higher risk for PTSD relating to assaultive violence exposure is consistent with previous reports from the Detroit community[6, 7] and other studies of events relating to assaultive violence[4, 69].

This study included some limitations. Due to overlaps in participant-reported trauma exposures in different waves of the DNHS, it is possible that our relatively small sample size did not have sufficient statistical power to detect significant differences in methylation at cg17057218, cg22324981, or cg04755409 across the trauma types.

Nevertheless, we were able to detect small, but significant, differences in methylation levels at cg22324981 and/or cg17057218 associated with each of the five trauma types. Second, our assessment of 5-mC at one time-point (W1/W2), combined with an assessment of lifetime PTSD rather than PTSD onset, leaves us unable to conclude whether the observed differences in 5-mC precede PTSD or trauma exposure; however, because we prospectively assessed PTSD at wave 3, we *can* say that the 5-mC differences associated with trauma exposures reported at W1/W2 are associated with subsequent psychopathology reported at W3, enhancing the causal framework of our analyses. Third, our study did not adjust for the number of lifetime trauma types, which has been previously identified as a significant predictor of PTSD and symptom presentation[20, 70]. Lastly, our study focused on trauma types for which PTSD was assessed, however, this does not exclude the possibility that participants can have PTSD from other trauma types that were not selected as the worst or random event in the DNHS.

Despite these limitations, our study is strengthened by its analytical methods, which consisted of identifying PTSD-relevant CpG sites, that were then interrogated using path analysis GSEM, followed by a mediational assessment method for binary outcomes (W3 lifetime PTSD) that enabled us to assess the DE, IDE, and TE for various trauma types. Whereas other methylation studies have primarily focused on early childhood traumas or individual traumatic events, we assessed the relative importance of different trauma types on *NFATC1* 5-mC and differential risk of prospective PTSD. Furthermore, we attempted to identify some preliminary and potential biological relevance of the mediating CpG sites as well as placing *NFATC1*‘s expression in the context of brain tissue. In sum, our results indicate a relationship between some trauma types (assaultive violence, other injury or shocking experience, learning of trauma to a loved one, and other extraordinarily stressful situation or event), *NFATC1* 5-mC, and prospective risk of PTSD. Future studies are needed to assess and corroborate the importance of *NFATC1* 5-mC in the overall context of trauma exposure and PTSD psychopathology, as well as its relation to other genes, ideally in larger cohorts. Moreover, more work is needed to untangle the biological mechanisms underlying the complex relationship between different trauma types and the differential risk for PTSD.

## Data Availability

All data produced in the present study are available upon reasonable request to the authors.

## Acknowledgments

This work was supported by the National Institutes of Health grants R01DA022720 R01DA022720-S1, RC1MH088283, R01MD011728, and R01MD011728-S2. We are very grateful to the study participants, staff, and volunteers of the Detroit Neighborhood Health Study for their time and contributions to the study.

